# Access to Allogeneic Cell Transplantation Based on Donor Search Prognosis: An Interventional Trial

**DOI:** 10.1101/2024.09.16.24313494

**Authors:** Jason G. Dehn, Brent Logan, Bronwen E. Shaw, Steven Devine, Stefan O. Ciurea, Mary Horowitz, Naya He, Iskra Pusic, Samer A. Srour, Sally Arai, Mark Juckett, Joseph Uberti, LaQuisa Hill, Sumithira Vasu, William J. Hogan, Brandon Hayes-Lattin, Peter Westervelt, Asad Bashey, Nosha Farhadfar, Michael R. Grunwald, Eric Leifer, Heather Symons, Ayman Saad, Jenny Vogel, Connor Erickson, Kelly Buck, Stephanie J. Lee, Joseph Pidala

## Abstract

**Importance:** Patients requiring allogeneic hematopoietic cell transplantation have variable likelihoods of identifying an 8/8 HLA-matched unrelated donor. A Search Prognosis calculator can estimate the likelihood.

**Objective:** To determine if using a search algorithm based on donor search prognosis can result in similar incidence of transplant between patients Very Likely (>90%) vs Very Unlikely (<10%) to have a matched unrelated donor.

**Design:** This interventional trial utilized a Search Prognosis-based biologic assignment algorithm to guide donor selection. Trial enrollment from June 13, 2019-May 13, 2022; analysis of data as of September 7, 2023 with median follow-up post-evaluability of 14.5 months.

**Settings:** National multi-center Blood and Marrow Transplantation Clinical Trials Network 1702 study of US participating transplant centers.

**Participants:** Acute myeloid and lymphoid leukemias, myelodysplastic syndrome, Hodgkin’s and non-Hodgkin’s lymphomas, severe aplastic anemia, and sickle cell disease patients referred to participating transplant centers were invited to participate. 2225 patients were enrolled and 1751 were declared evaluable for this study. Patients were declared evaluable once it was determined no suitable HLA-matched related donor was available.

**Intervention:** Patients assigned to the Very Likely arm were to proceed with matched unrelated donor, while Very Unlikely were to utilize alternative donors. A third stratum, Less Likely (∼25%) to find a matched unrelated donor, were observed under standard center practices, but were not part of the primary objective.

**Main Outcome:** Cumulative incidence of transplantation by Search Prognosis arm

**Results:** Evaluable patients included 1751 of which 413 (24%) were from racial/ethnic minorities. Search prognosis was 958 (55%) Very Likely, 517 (30%) Less Likely and 276 (16%) Very Unlikely. 1171 (67%) received HCT, 384 (22%) died without HCT, and 196 (11%) remained alive without HCT. Among the 1,234 patients, the adjusted cumulative incidence (95% CI) of HCT at 6-months was 59.8% (56.7-62.8) in the Very Likely group versus 52.3% (46.1-58.5) in the Very Unlikely (P=0.113).

**Conclusions:** A prospective Search Prognosis-based algorithm can be effectively implemented in a national multicenter clinical trial. This approach resulted in rapid alternative donor identification and comparable rates of HCT in patients Very Likely and Very Unlikely to find a matched unrelated donor.

**Trial Registration:** NCT#03904134

## INTRODUCTION

Allogeneic hematopoietic cell transplantation (HCT) is a potentially curative therapy for hematological malignancies and non-malignant diseases.^1,2^ Typically the first choice of a donor is an HLA matched related donor; however, only approximately 30% of patients have one identified (range 13-50% depending on race and age).^3,4^ Generally, the second choice is a matched unrelated donor (MUD) (8/8 high resolution allele match at HLA-A, -B, -C, and - DRB1), available from worldwide donor registries, with HCT outcomes similar to matched related donor transplants.^5–10^ Despite a large donor inventory (www.wmda.info), the likelihood of finding a MUD differs based on patient race and ethnicity. In the United States, patients of White-European descent have 8/8 MUD match rates of 77% compared to African-Black rates of 23%, and rates around 45% in Hispanic and Asian patients.^11,12^ Therefore, patients from racial/ethnic minorities are more likely to receive HLA mismatched HCTs, experience delays in time to transplantation,^13^ and be less likely to receive HCT.^14,15^ Prolonged MUD searches may result in an increased risk of pretransplant relapse/progression of disease.

When fully matched donors are not available, most HCT centers consider using alternative donors, including mismatched unrelated donors (MMUD), umbilical cord blood (UCB), or related haploidentical donors (Haplo), as outcomes from HCT using these donor types have improved.^16–20^ However, the ideal donor search and selection strategy for a given patient is uncertain. There is high variability in alternative donor search practices with some delaying the search for an alternative donor until an exhaustive search for a matched donor proves futile. The National Marrow Donor Program^®^ (NMDP) developed a Search Prognosis calculator, which uses HLA haplotype frequency data along with a patient’s race and ethnicity to predict whether the patient is likely or not to have an 8/8 MUD.^21^ The resulting classification predicts whether a patient is Very Likely (>90%), Less Likely (∼25%) or Very Unlikely (<10%) to have a MUD identified. This allows early identification of the best donor selection strategy. Here we report the results of the National Institutes of Health funded Blood and Marrow Transplant Clinical Trial Network (BMT CTN) 1702 sub-study which utilized the Search Prognosis score to assign patients to arms with different donor search and selection strategies, with the objective of providing a rapid and efficient path to HCT for all patients.

## METHODS

In this study we performed a pre-planned secondary analysis from the multicenter BMT CTN 1702 (protocol available at www.bmtctn.net). The primary objective of BMT CTN 1702 was to estimate and compare the overall survival between two arms: patients who were Very Likely to find a MUD and, per protocol, pursued a MUD search versus those who are Very Unlikely to find a MUD and, per protocol, immediately pursued an alternative donor early. Patients categorized as Less Likely to find MUD were not part of the primary objective. Their donor search strategy was not specified by the protocol and observational data were analyzed in descriptive analyses. Objectives of the current study were to compare the incidence of and time to transplantation in the two groups and to identify factors delaying or precluding HCT. Eligibility criteria included patients of any age with acute myeloid leukemia (AML), acute lymphoblastic leukemia (ALL), myelodysplastic syndrome (MDS), non-Hodgkin’s lymphoma (NHL), Hodgkin’s lymphoma (HL), severe aplastic anemia (SAA) or sickle cell disease (SCD) who were considered suitable HCT candidates. Additionally, centers must have confirmed their intention both to follow the protocol-recommended search algorithm and to perform the HCT within the next six months. Consented patients became evaluable once it was determined no suitable HLA-matched related donor was available. Exclusion criteria were patients with prior allogeneic HCT or previous formal unrelated donor search.

Once enrolled and HLA typing and race/ethnicity data submitted, the NMDP provided centers unrelated donor search results and Search Prognosis: MUD Very Likely (>90%), MUD Less Likely (∼25%), or MUD Very Unlikely (<10%). Based on the Search Prognosis, the trial’s biologic assignment^22^ algorithm advised centers that the Very Likely group should proceed with 8/8 URD search and HCT and the Very Unlikely group should proceed with alternative donor HCT based on pre-specified rank priority (e.g. UCB, Haplo, MMUD). The search strategy for the Less Likely group was left to institutional practice. Monthly center reporting was submitted including any change to the HCT timeframe or alternative donor priority. Based on prior publications, the projected biological arm assignment was 44% Very Likely, 41% Less Likely, and 15% Very Unlikely.^21^

Enrollment began June 13, 2019 and ended May 13, 2022 with 53 transplant centers activated. The prespecified objectives of this secondary analysis were (1) to estimate and compare the cumulative incidence of receiving a HCT when using an upfront donor Search Prognosis score, and (2) to describe the barriers to achieving HCT based on Search Prognosis. An HCT delay was defined as a change in the target patient transplant date; an HCT search cancellation was defined as the discontinuation of active pursuit of transplant. Patient reported race and ethnicity data was collected in accordance with NIH guidelines and followed Office of Management and Budget standards. This analysis includes follow up data as of September 7, 2023 and was approved by the Data and Safety Monitoring Board for analysis prior to the primary endpoints being reached. The research protocol was approved by the NMDP IRB. All study subjects (minor’s parents/guardians) provided written informed consent. The trial is registered in clinicaltrials.org with NCT03904134.

## Statistical Analysis

The sample size was powered on analysis of the primary study objective, overall survival, which will be reported later; details are available in the protocol. The primary analysis population is all enrolled patients who are declared evaluable, and who were biologically assigned to the Very Likely or Very Unlikely group. The Less Likely group was analyzed separately in a descriptive analysis. The secondary outcomes in this report were analyzed according to the donor Search Prognosis assignment regardless of what donor was actually prioritized or used as an intention-to-treat analysis and were measured from the time the patient was deemed evaluable. Patient and search characteristics (including HCT delays or cancellations and their reasons) were described by donor Search Prognosis category using frequencies and percents for categorical variables and median (range) for continuous variables. Categorical variables were compared between arms using the chi-square test or Fisher’s exact test as appropriate, while continuous variables were compared using the Kruskal-Wallis test. Cumulative incidences of HCT were described using the Aalen-Johansen estimator^23^, treating death prior to HCT as a competing risk. Cumulative incidences of HCT were compared between Very Likely and Very Unlikely groups using a Fine-Gray model^24^, adjusting for the following pre-specified characteristics: age, sex, race, ethnicity, performance score, time from enrollment to evaluability, disease and disease status. Adjusted cumulative incidence curves were also generated.^25^ The statistical analyses were performed with the use of SAS software, version 9.4 or R V3.6.3 and conducted prior to the primary study endpoint comparison as described in a formal baseline statistical analysis plan for the protocol.

## RESULTS

### Population

Patient baseline characteristics are shown in Table 1 by Search Prognosis assignment. The total study enrollment of 2225 patients resulted in a final analysis cohort of 1751 evaluable patients enrolled from 47 total HCT centers (of 53 centers activated, 2 did not have an evaluable patient; 4 did not enroll); 958 (55%) patients were Very Likely, 517 (30%) Less Likely, and 276 (16%) Very Unlikely. Overall patient median age was 59 years, 50% of patients had AML, and 46% had a Karnofsky performance score >=90%. Patient race and ethnicity was 0.6% American Indian/Alaskan Native, 4.7% Asian/Native Hawaiian/Pacific Islander, 7.5% Black/African American, 10.3% Hispanic White, 73.2% Non-Hispanic White, and 3.6% other/multiple race/missing. Median (range) follow-up was 14.5 (0.1-43.7) months.

**Table 1:** Patient Characteristics at Trial Enrollment by Search Prognosis

| Characteristic | Very<br>Likely | Less<br>Likely | Very<br>Unlikely | P<br>Value <sup>a</sup> | Total |
| --- | --- | --- | --- | --- | --- |
| **No. of patients (%)** | 958 (55) | 517 (30) | 276 (16) |  | 1751 |
| **No. of centers** | 43 | 45 | 40 |  | 47 |
| **Sex - no. (%)** |  |  |  | 0.90 |  |
| Male | 539 (56) | 286 (55) | 157 (57) |  | 982 (56) |
| Female | 419 (44) | 231 (45) | 119 (43) |  | 769 (44) |
| **Age at evaluable,**<br>**years - no. (%)** |  |  |  | <.01 |  |
| Median (min-max) | 61.2 (0.7-81.3) | 57.8 (0.6-79.7) | 56.4 (2.6-76.4) |  | 59.4 (0.6-81.3) |
| <=10 | 17 (2) | 16 (3) | 8 (3) |  | 41 (2) |
| 11-20 | 21 (2) | 28 (5) | 13 (5) |  | 62 (4) |
| 21-30 | 61 (6) | 49 (10) | 25 (9) |  | 135 (8) |
| 31-40 | 61 (6) | 41 (8) | 28 (10) |  | 130 (7) |
| 41-50 | 99 (10) | 48 (9) | 32 (12) |  | 179 (10) |
| 51-60 | 190 (20) | 104 (20) | 56 (20) |  | 350 (20) |
| 61-70 | 351 (37) | 161 (31) | 84 (30) |  | 596 (34) |
| >70 | 158 (17) | 70 (14) | 30 (11) |  | 258 (15) |
| **KPS at evaluability**<br>**- no. (%)** |  |  |  | 0.32 |  |

| **Characteristic** | **Very**<br>**Likely** | **Less**<br>**Likely** | **Very**<br>**Unlikely** | **P**<br>**Value <sup>a</sup>** | **Total** |
| --- | --- | --- | --- | --- | --- |
| >=90 | 416 (43) | 245 (47) | 138 (50) |  | 799 (46) |
| 80 | 330 (34) | 160 (31) | 91 (33) |  | 581 (33) |
| <80 | 207 (22) | 108 (21) | 46 (17) |  | 361 (21) |
| Missing | 5 (1) | 4 (1) | 1 (<1) |  | 10 (1) |
| **Race/Ethnicity - no. (%)** |  |  |  | **<.01** |  |
| American Indian /Alaska<br>Native | 5 (0.5) | 3 (0.6) | 2 (0.7) |  | 10 (0.6) |
| Asian/Native<br>Hawaiian/Pacific Islander | 18 (2) | 37 (7) | 28 (10) |  | 83 (5) |
| Black /African American | 16 (2) | 76 (15) | 40 (15) |  | 132 (8) |
| Hispanic White | 50 (5) | 88 (17) | 43 (16) |  | 181 (10) |
| Non-Hispanic White | 846 (88) | 291 (56) | 145 (53) |  | 1282 (73) |
| Other/Multiple Race | 1 (<1) | 2 (<1) | 4 (1) |  | 7 (<1) |
| Unknown/Missing | 22 (2) | 20 (4) | 14 (5) |  | 56 (3) |
| **Disease - no. (%)** |  |  |  | **<.01** |  |
| AML | 496 (52) | 250 (48) | 130 (47) |  | 876 (50) |
| ALL | 146 (15) | 97 (19) | 62 (23) |  | 305 (17) |
| MDS | 246 (26) | 116 (22) | 55 (20) |  | 417 (24) |
| HL | 6 (1) | 5 (1) | 1 (<1) |  | 12 (1) |
| NHL | 32 (3) | 28 (5) | 20 (7) |  | 80 (5) |
| Severe aplastic anemia | 25 (3) | 11 (2) | 5 (2) |  | 41 (2) |
| Sickle cell | 3 (<1) | 2 (<1) | 2 (1) |  | 7 (<1) |
| Other acute leukemia | 4 (<1) | 8 (2) | 1 (<1) |  | 13 (1) |
| **Target time to start of**<br>**conditioning - no. (%)** |  |  |  | 0.91 |  |
| Less than 6 weeks | 116 (12) | 61 (12) | 35 (13) |  | 212 (12) |
| Over 6 weeks up to 12<br>weeks | 492 (51) | 281 (54) | 145 (53) |  | 918 (52) |
| Over 12 weeks up to 6<br>months | 349 (36) | 174 (34) | 96 (35) |  | 619 (35) |
| Missing | 1 (<1) | 1 (<1) | 0 (0) |  | 2 (<1) |
| **Disease status,**<br>**AML/ALL/OAL - no. (%)** |  |  |  | 0.38 |  |
| CR1/CR2 | 211 (33) | 116 (33) | 73 (38) |  | 400 (34) |
| CR3+/PIF/Rel | 101 (16) | 52 (15) | 30 (16) |  | 183 (15) |
| No Treatment/ Currently<br>undergoing induction<br>treatment/ Unknown | 332 (51) | 183 (52) | 90 (47) |  | 605 (51) |
| Missing | 2 (<1) | 4 (1) | 0 (0) |  | 6 (1) |
| **Disease status, MDS - no.**<br>**(%)** |  |  |  | 0.78 |  |
| CR/Hi | 21 (9) | 12 (10) | 7 (13) |  | 40 (10) |
| NR/SD/Prog from HI/Rel<br>from CR | 89 (36) | 42 (36) | 16 (29) |  | 147 (35) |
| Not assessed | 136 (55) | 62 (53) | 32 (58) |  | 230 (55) |
| **Disease status, HL/NHL -**<br>**no. (%)** |  |  |  | **0.47** |  |
| Treatment responsive | 9 (24) | 11 (33) | 8 (38) |  | 28 (30) |
| Not Treatment responsive | 29 (76) | 22 (67) | 13 (62) |  | 64 (70) |
| **The first preferred**<br>**alternative donor -**<br>**no.(%)** |  |  |  | **0.02** |  |
| None | 33 (3) | 5 (1) | 3 (1) |  | 41 (2) |
| Mismatched unrelated | 274 (29) | 153 (30) | 81 (29) |  | 508 (29) |
| Haploidentical related | 597 (62) | 318 (62) | 174 (63) |  | 1089 (62) |
| Umbilical Cord blood | 54 (6) | 39 (8) | 18 (7) |  | 111 (6) |
| Not reported | 0 (0) | 2 (<1) | 0 (0) |  | 2 (<1) |

| Characteristic | Very<br>Likely | Less<br>Likely | Very<br>Unlikely | P<br>Value <sup>a</sup> | Total |
| --- | --- | --- | --- | --- | --- |

### Donor Search and Selection

Reported by the HCT center at each patient enrollment, first preference of alternative donor was 62% Haplo, 29% MMUD, and 6% UCB in the total study cohort. Figure 3 shows the center-specified alternative donor source priority by year of study enrollment, showing increased preference for MMUD and reduced preference for Haplo and UCB in the later years of the trial.

### Incidence and Time to Transplant

Overall, 67% of patients received HCT, 22% died without HCT and 11% were alive without HCT at study end. Among patients who received HCT, donors used for patients in Very Likely arm were MUD (94%), Haplo (4%), MMUD (1%) and UCB (1%). Donors used in the Less Likely arm were MUD (38%), Haplo (37%), MMUD (20%), and UCB (6%). Donors used in the Very Unlikely arm were MUD (9%), Haplo (61%), MMUD (22%), and UCB (8%). Figure 1a shows the unadjusted cumulative incidence of HCT by Search Prognosis assignment. Among the 1,234 patients in the Very Likely and Very Unlikely arms, cumulative incidences of HCT adjusted for patient and donor search related variables showed no statistically significant difference (Figure 1b, P=0.113). Pre-HCT death rates and median time from enrollment to evaluability were not statistically different between Search Prognosis groups. HCT incidences at six-months were 59.8% in Very Likely vs 52.3% in Very Unlikely arms and at two-years 70.1% and 64.3%, respectively. The corresponding median (range) times to HCT were 3.3 (0.1-39.7) months and 3.3 (0.5-35.4) months.

**Figure 1:**
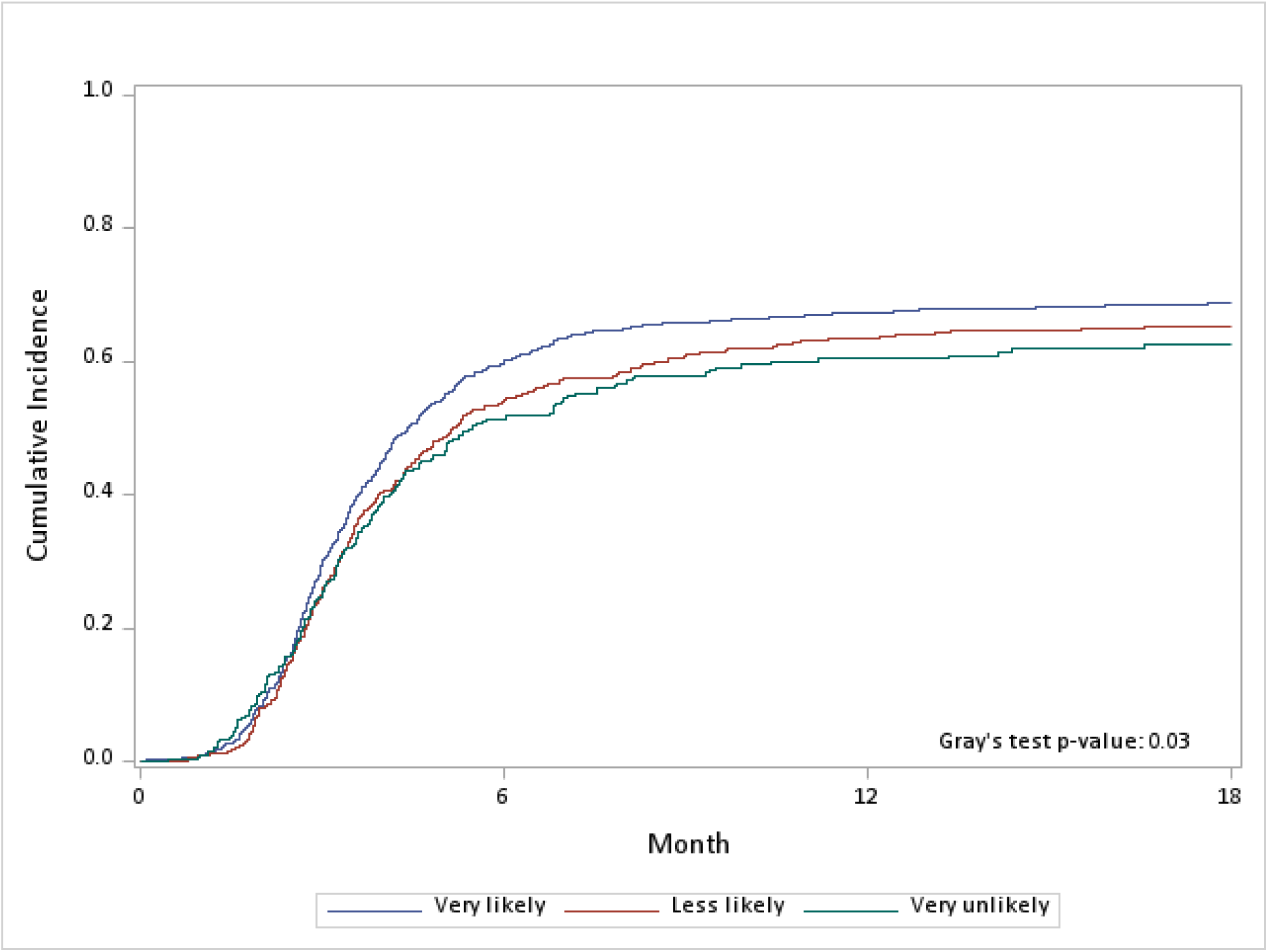
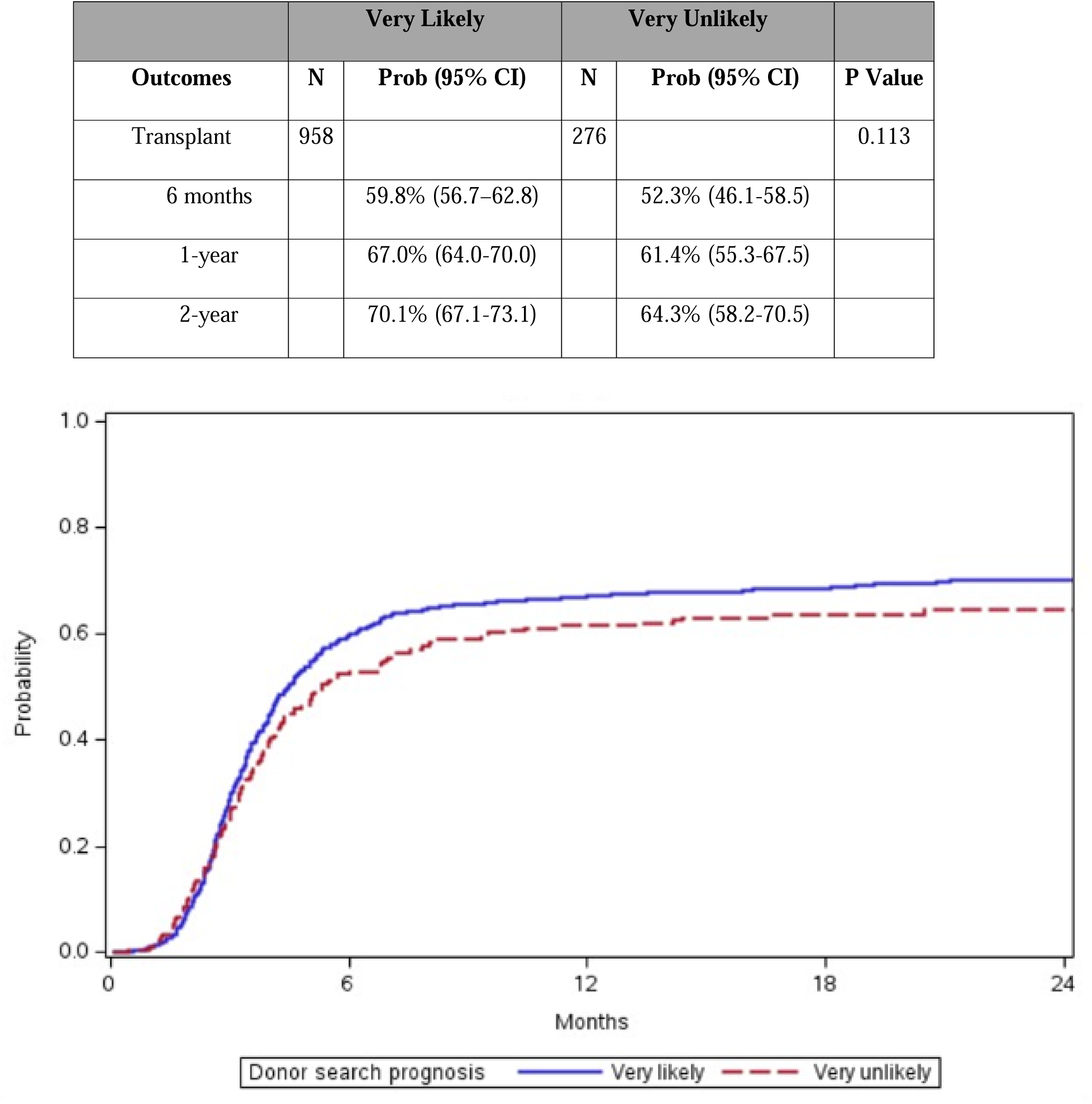
Cumulative Incidence of Transplant. **a) Unadjusted Cumulative Incidence of Transplant by Search Prognosis Arm** Graph shows the unadjusted cumulative incidence of hematopoietic cell transplant over time from patient evaluability. **b) Adjusted Cumulative Incidence of Transplant by Donor Search Prognosis Arm** Graph shows the adjusted cumulative incidence of hematopoietic cell transplant by Search Prognosis arm over time from patient evaluability; Very Likely and Very Unlikely.

Table 2 shows the multivariate analysis between Very Likely and Very Unlikely group. Older patient age, lower Karnofsky performance score, patient disease other than AML and disease status other than complete remission were significantly associated with lower rate of HCT.

**Table 2:**
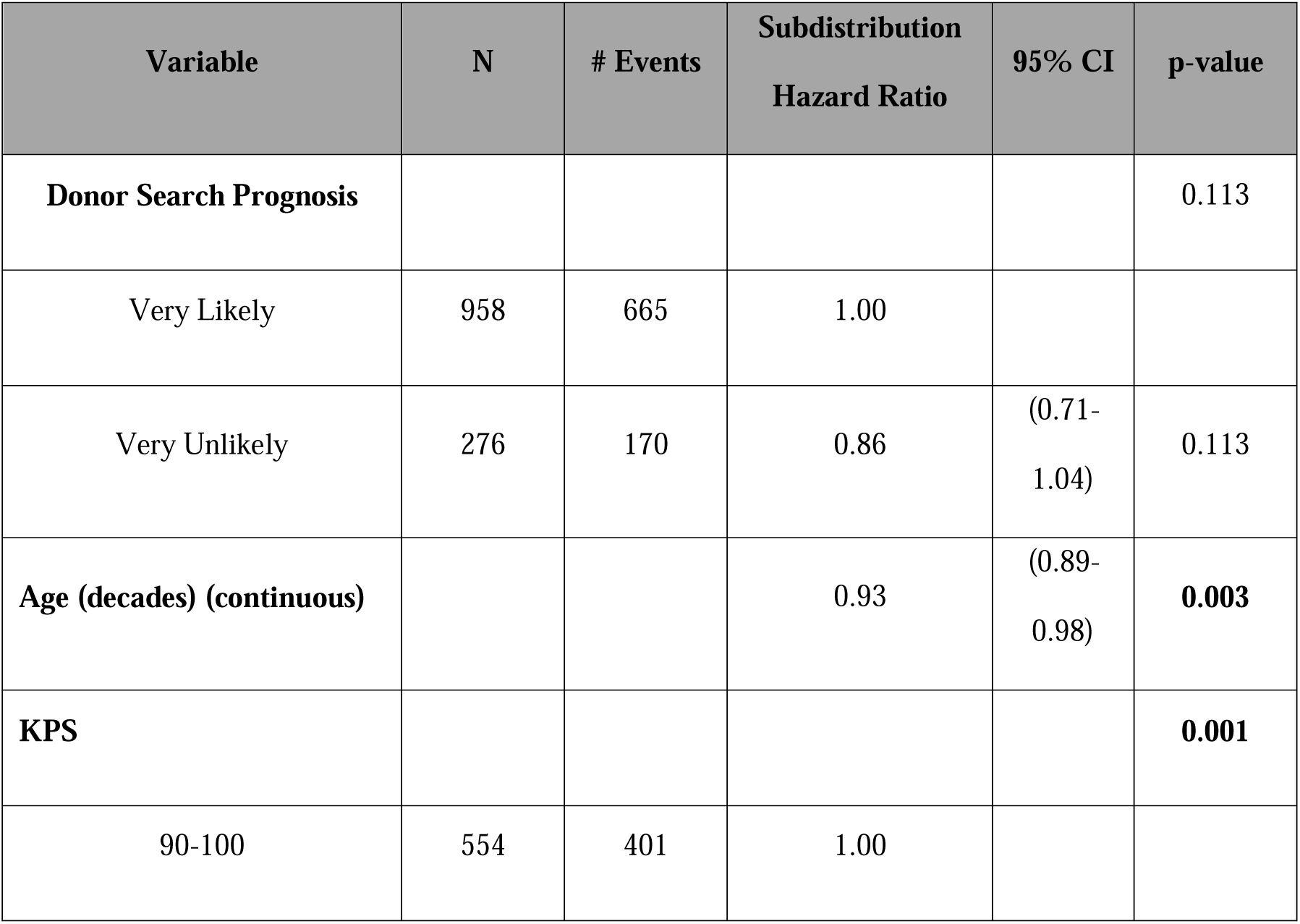
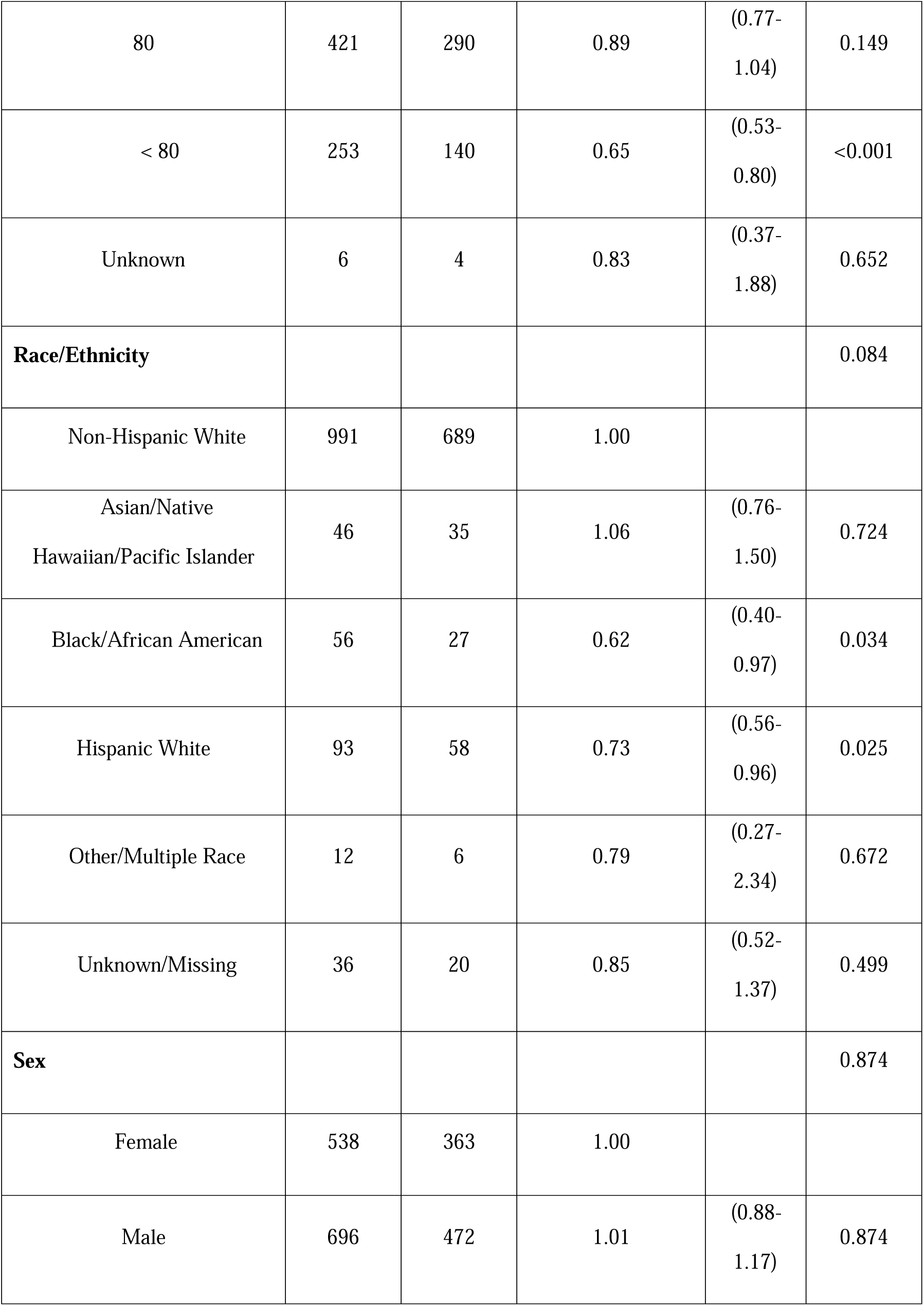
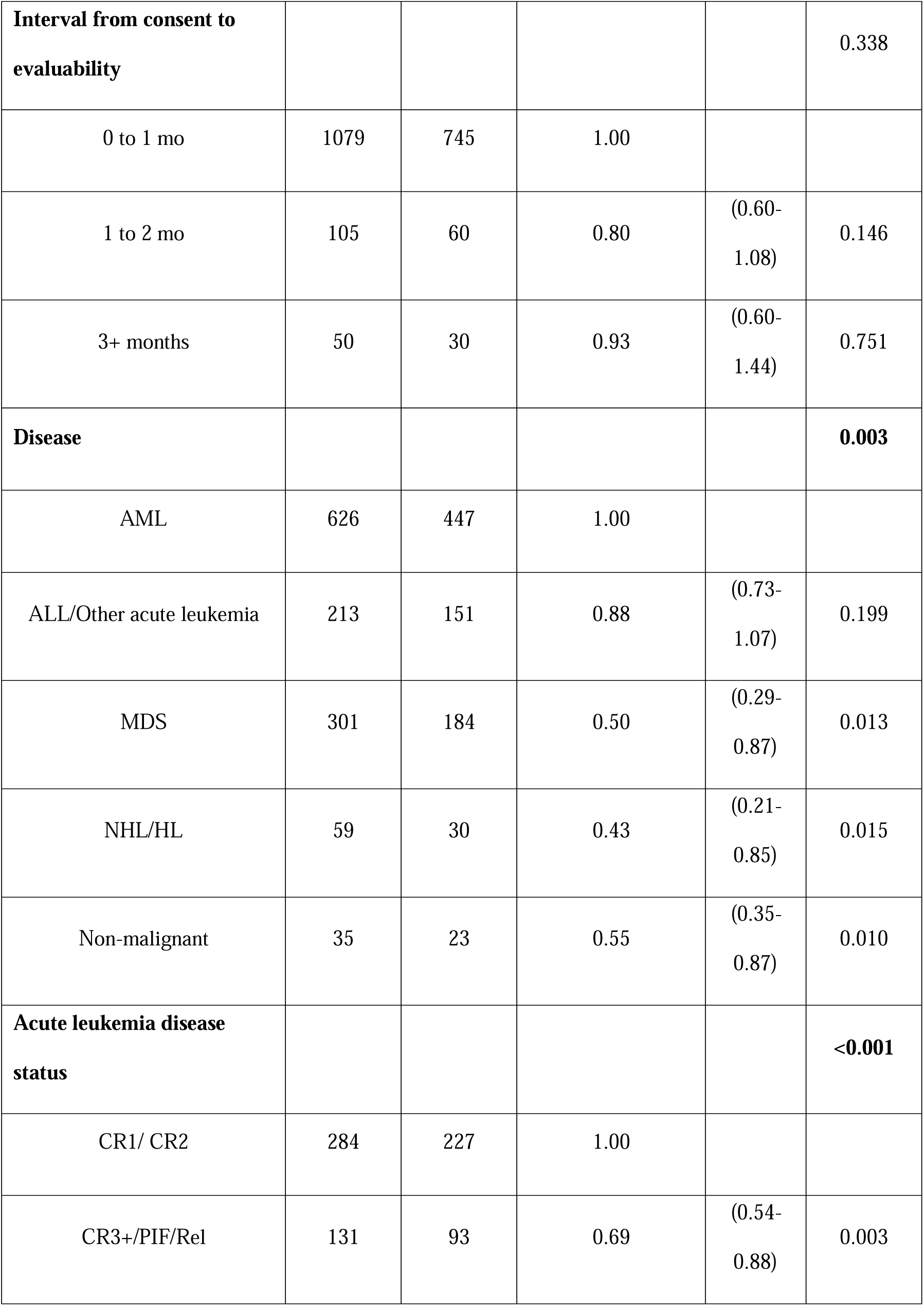
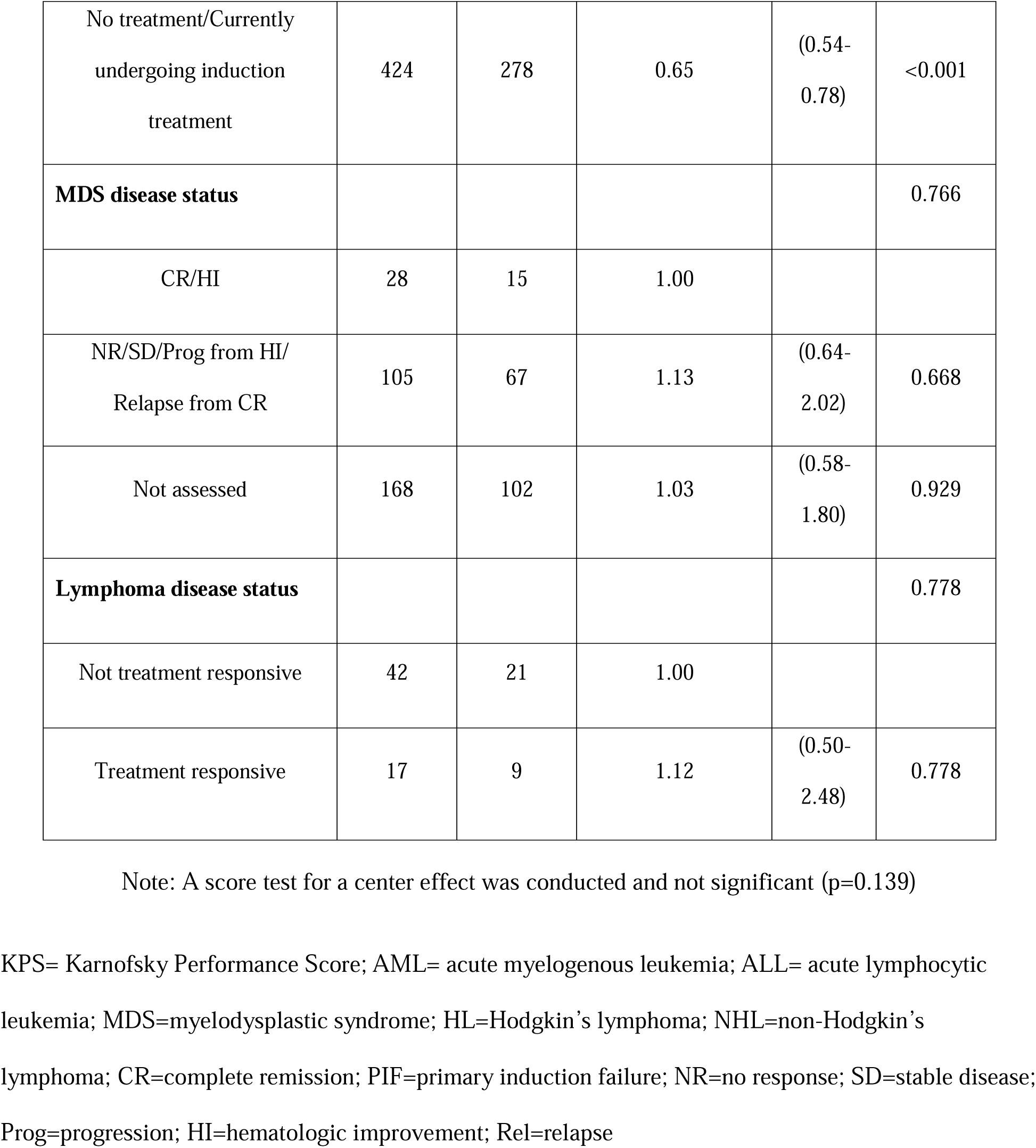
Multivariate Fine-Gray Analysis of Transplantation in the Primary Analysis Population

In a subset analysis of patients who were transplanted in this study (Figure 2), median time from evaluability to HCT by donor source did not reach statistical significance ranging from 3.8 months for UCB, 3.6 months for MMUD, 3.4 months for MUD to 3.2 months for Haplo (P=0.06).

**Figure 2:**
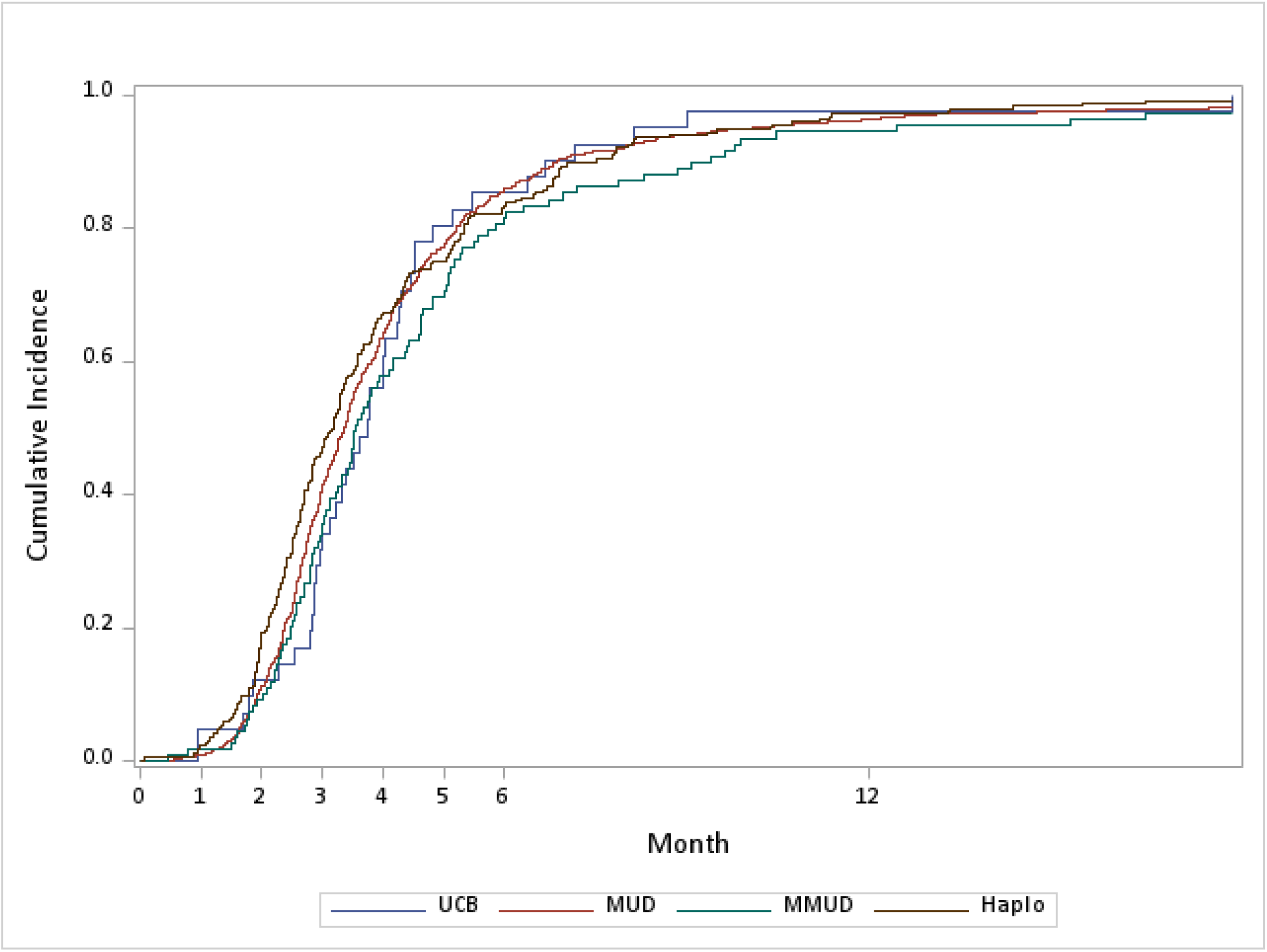
Time from Evaluability to Transplant Among Transplanted Patients. Graph shows the cumulative incidence of transplant by infused donor type for patients who reach transplant. UCB=umbilical cord blood; MUD= matched unrelated donor; MMUD=mismatched unrelated donor; Haplo=haploidentical related donor

**Figure 3:**
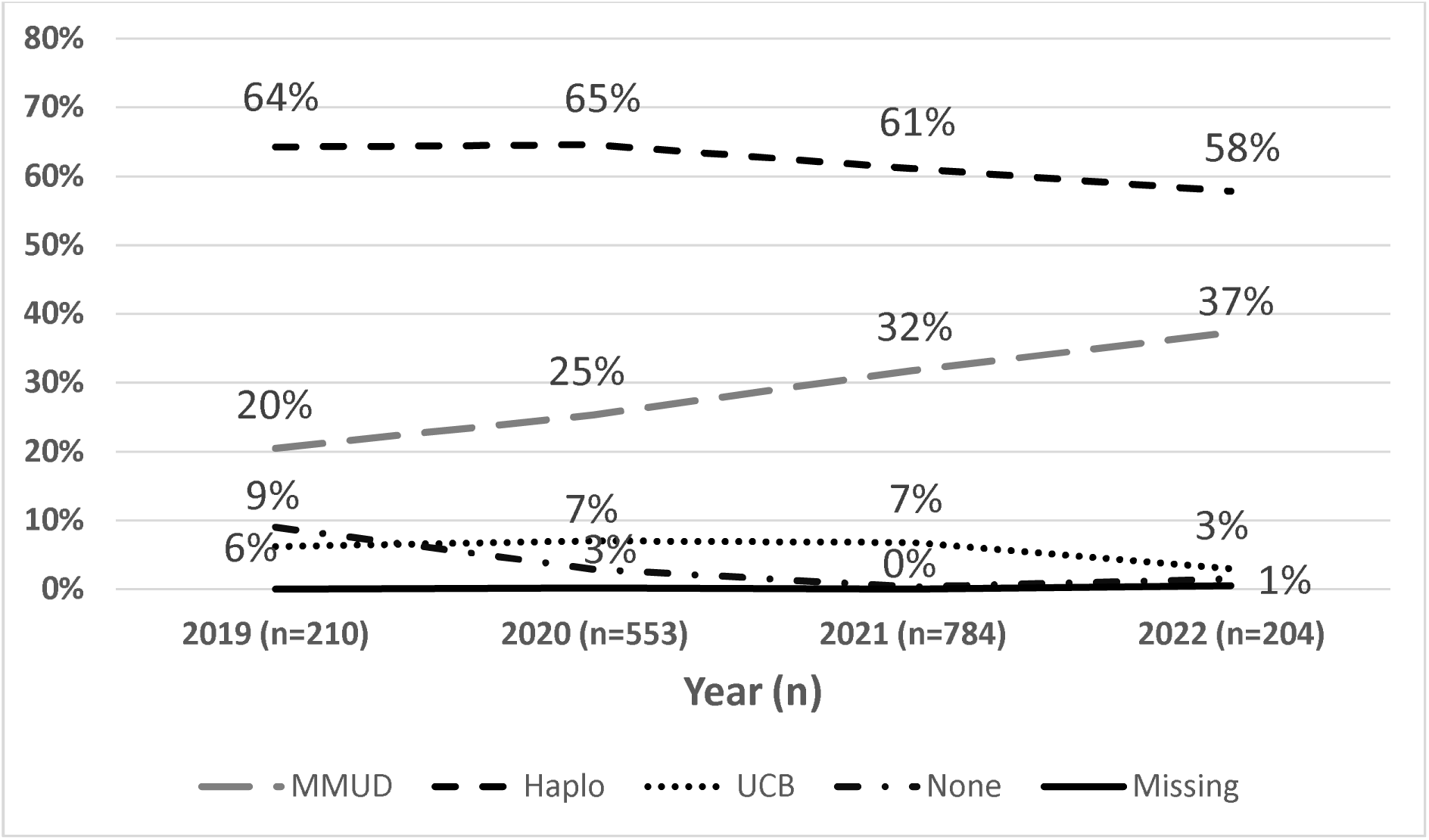
First Preference of Donor Cell Source by Year of Enrollment. Graph shows the reported first ranked donor cell source at enrollment by year. Mismatched unrelated donor (MMUD), haploidentical related (Haplo) and Umbilical Cord Blood (UCB), No donor cell alternatives reported (None) and Missing are shown.

### Delays/Cancellations

The reasons for first delay or search cancellation by Search Prognosis group are shown in Supplementary Table S2 and S3, respectively. In the total study population, 59% of delays and cancellations were due to patient poor health and around 9% percent of patients reported excellent patient disease response to nontransplant therapy. Fourteen percent of delays and 2% of cancellations were due to donor issues.

Primary delay (p=0.03) and cancellation (p=<0.01) reason differences by Search Prognosis group were statistically significant. Delays showed patient poor health was increased in Very Likely group and excellent patient response increased in the Very Unlikely arm. Cancellation data showed higher proportion of patient poor health (64%) for the Very Likely group compared to 45% in the Very Unlikely arm. Cancellation reason of patient preference was highest in Very Unlikely arm, with 20% of the cancellations reported.

## DISCUSSION

This study demonstrates that using an early Search Prognosis to guide donor selection, with immediate pursuit of alternative donors when a MUD is unlikely, results in comparable rates of receiving HCT regardless of other baseline prognostic factors. Patients in the Very Likely group identified and proceeded with a MUD HCT in the great majority of cases. Conversely, in the Very Unlikely group, HCT using an alternative donor occurred in most cases. Incidence and time to HCT was comparable between the Search Prognosis arms. These are exciting developments given the low rates of 8/8 MUD available for patients of racial and ethnic minorities compared to their non-Hispanic White counterparts, driven by inadequate representation within worldwide registries of donors of non-European ancestry, compounded by higher HLA diversity in some groups, particularly those of African descent. Requirements for stringent matching in this setting make searching for a MUD a near-futile exercise in some groups.

Remarkably this approach was accomplished in a large study of 47 HCT centers that assigned patients to treatment arms using this simple and timely Search Prognosis tool. The trial achieved notable racial and ethnic representation, with 24% of participants from groups other than non-Hispanic Whites which is comparable to 23% of patients overall with formal search activity through the NMDP in the same study period from the study participating centers, matched for patient disease criteria (Supplementary Table S1). In the current treatment era when almost all patients have at least one available donor, improving patient HCT outcomes by enabling early identification of patients with a very low likelihood of finding a MUD, and immediately switching to available alternative donor sources is critical to ensuring the maximum number of people benefit from the therapy. Over the duration of the study, transplant center ranking preference evolved for alternative donor sources. Recent accumulation of published data suggesting impressive outcomes using post-transplant cyclophosphamide in the MMUD setting may have contributed to increased consideration of this alternative.^16,17^

Prior research and development of the Search Prognosis Score was used to predict how the algorithm would perform in this clinical trial. Biological assignment for the study was projected to result in 15% in the Very Unlikely group (close to the actual result of 16%) and 44% in the Very Likely group (actual = 55%). Additionally, the algorithm performed very well in predicting the use of specific donors, as those transplanted in the Very Likely arm underwent MUD HCT 94% of the time (predicted at >90%), and those in the Very Unlikely group used alternative cell sources 91% of the time (predicted at >90%). This suggests that for all HCT centers embarking on a donor search, the Search Prognosis algorithm is a useful tool to guide search strategy. This is easily implementable as the Search Prognosis calculator used in this study requires only the patient’s race/ethnicity and HLA type to provide the prediction. The tool is freely available at https://search-prognosis.b12x.org/.

Based on the results of this study, early consideration of alternative donor sources can assist in equalizing opportunities for HCT with a standardized approach, while those with a Very Likely score are not penalized, in terms of the probability or timing of HCT, by pursuing a fully matched donor. The outcomes of this sub-study await some application guidance pending analysis of patient clinical outcome differences, if any, between the Very Likely vs Very Unlikely arms, the subject of the BMT CTN 1702 primary analysis. Regardless, the availability of a simple and effective tool to standardly guide centers early in the donor search process is critical.

Importantly, about one-third of patients did not reach HCT due to several non-donor-related barriers. These included poor health status (primarily disease factors and newly developed co-morbidities) of the patient that did not allow for timely or full progression to HCT, which was a major factor in both HCT delays and cancellations. Additionally, about 9% of patients were felt to no longer require urgent HCT (or perhaps HCT at all) based on excellent response to other therapy, predominantly patients in first complete remission. Another common reason for search cancellations was the preference of the patient to discontinue the donor search, which was more common in the Very Unlikely group. Unfortunately, limited details were available as to the reasons for those decisions. Donor issues were a minor factor contribution to HCT delay or cancellation. Further investigation into strategies focused on maintaining or improving patient clinical status to allow for HCT and mitigation of potential development of co-morbidities during treatment are needed but will have to contend with the complexity of attempting to control aggressive diseases while minimizing toxicity.

We acknowledge several limitations to this study. The cohort was predominantly patients of older age (and limited pediatric representation). Some demographic differences between patients in the Search Prognosis arms may not fully be accounted for in covariate adjusted modeling, including social determinants of health, some clinical/disease related aspects, or with all transplant center specific differences in the path to HCT. Finally, although this study measures access of those engaged with an active HCT center, it does not address the disparities of access that exist at the community level where patients must first be connected to a transplant center before donor considerations are addressed.

Our study shows a major advance in overcoming current limitations with equitable and timely access to HCT for patients likely and unlikely to have a MUD. The latter group is notably enriched with diverse race/ethnic patients who are at an unacceptable disadvantage in receiving HCT if the focus is on a fully matched donor. However, MUD and alternative donor sources are accessible with a probability of HCT in a similar time frame with support from a Search Prognosis calculator-based strategy allowing the process to move in a timely manner. Overall, our study shows exciting promise for guiding clinical decisions and extending the cure rates in the current era where finding a donor for every patient is a reality.

## Data Availability

Data will be submitted by the Blood and Marrow Transplant Clinical Trials Network (BMT CTN) to the National Heart, Lung, and Blood Institute (NHLBI) Data Repository within a month of peer-reviewed primary study results publication and will be accessible via standard Repository processes.

https://bmtctn.net/bmt-ctn-studies

## ACKNOWLEDGMENTS

Support for this study was provided by grants #U10HL069294 and #U24HL138660 to the Blood and Marrow Transplant Clinical Trials Network from the National Heart, Lung, and Blood Institute and the National Cancer Institute. The content is solely the responsibility of the authors and does not necessarily represent the official views of the NIH. The CIBMTR registry is supported primarily by the U24-CA76518 from the National Cancer Institute, the National Heart, Lung, and Blood Institute, and the National Institute of Allergy and Infectious Diseases and from HHSH234200637015C (HRSA/DHHS) to the Center for International Blood and Marrow Transplant Research.

## AUTHORSHIP CONTRIBUTIONS

J.D. and J.P. wrote the paper. B.L. and N.H performed the statistical analyses.

All authors approved the manuscript and submission.

## CONFLICT-OF-INTEREST DISCLOSURE

S.J.L-Board of Directors, nmdp (uncompensated)

A.S.-Consulting Fees: Sanofi and Therakos

## Notes

### Clinical Trial

NCT03904134

### Author Declarations

The IRB of NMDP gave ethnical approval for this work.

