## Supplemental Table S1, S2, S3 for "Access to Allogeneic Cell Transplantation Based on Donor Search Prognosis: An Interventional Trial"

**Supplementary Appendix**

**Contents**

Supplementary Table S1	Page 3

Supplementary Table S2	Page 4

Supplementary Table S3	Page 5

**Supplementary Table S1:** Race and Ethnic Clinical Trial Enrollment Representation compared to all Patients matched for Disease and Study Transplant Centers: June 1, 2019 – August 31, 2022

| **Race and Ethnic Group** | **Study Enrollment** | **All Formal Unrelated Search Patients*** |
| --- | --- | --- |
| American Indian/Alaska Native | 0.6% | 0.4% |
| Asian/Native Hawaiian/Pacific Islander | 4.7% | 5.0% |
| Black/African American | 7.5% | 7.0% |
| Hispanic White | 10.3% | 9.4% |
| Non-Hispanic White | 73.2% | 68.0% |
| Other/Multiple Race | 0.4% | 1.0% |
| Unknown/Missing | 3.2% | 9.4% |

*NMDP formal unrelated search activity used internal NMDP data from accessed on March 11, 2024

**Supplementary Table S2:** Barriers to Hematopoietic Cell Transplantation for Overall Study Population: First HCT Delay

|  | **Very**  **Likely** | **Less Likely** | **Very Unlikely** | **P Value** | **Total** |
| --- | --- | --- | --- | --- | --- |
| **No. of patients** | 268 | 169 | 76 |  | 513 |
| **Primary reason for HCT delay**  **- no. (%)** |  |  |  | **0.03^a^** |  |
| Patient poor health | 169 (63.1) | 93 (55.0) | 40 (52.6) |  | 302 (58.9) |
| Excellent patient response | 21 (7.8) | 15 (8.9) | 11 (14.5) |  | 47 (9.2) |
| Donor | 34 (12.7) | 28 (16.6) | 8 (10.5) |  | 70 (13.6) |
| Transplant support | 6 (2.2) | 4 (2.4) | 3 (3.9) |  | 13 (2.5) |
| Patient preference | 7 (2.6) | 16 (9.5) | 8 (10.5) |  | 31 (6.0) |
| COVID | 21 (7.8) | 6 (3.6) | 2 (2.6) |  | 29 (5.7) |
| Other | 10 (3.7) | 6 (3.6) | 4 (5.3) |  | 20 (3.9) |
| Missing | 0 (0.0) | 1 (0.6) | 0 (0.0) |  | 1 (0.2) |

Hypothesis testing: ^a^ Pearson chi-square test

**Supplementary Table S3:** Barriers to Hematopoietic Cell Transplantation for Overall Study Population: HCT Search Cancellation

|  | **Very**  **Likely** | **Less Likely** | **Very Unlikely** | **P Value** | **Total** |
| --- | --- | --- | --- | --- | --- |
| **No. of patients** | 304 | 180 | 109 |  | 593 |
| **Primary reason for transplant cancellation - no. (%)** |  |  |  | **<.01^a^** |  |
| Patient poor health | 194 (63.8) | 106 (58.9) | 49 (45.0) |  | 349 (58.9) |
| Excellent patient response | 32 (10.5) | 13 (7.2) | 12 (11.0) |  | 57 (9.6) |
| Donor | 0 (0.0) | 9 (5.0) | 5 (4.6) |  | 14 (2.4) |
| Transplant support | 6 (2.0) | 6 (3.3) | 4 (3.7) |  | 16 (2.7) |
| Patient preference | 46 (15.1) | 29 (16.1) | 22 (20.2) |  | 97 (16.4) |
| COVID | 1 (0.3) | 1 (0.6) | 0 (0.0) |  | 2 (0.3) |
| Other | 25 (8.2) | 16 (8.9) | 17 (15.6) |  | 58 (9.8) |

Hypothesis testing: ^a^ Pearson chi-square test
